# Estimating the scale of COVID-19 Epidemic in the United States: Simulations Based on Air Traffic Directly from Wuhan, China

**DOI:** 10.1101/2020.03.06.20031880

**Authors:** Dalin Li, Jun Lv, Gregory Bowtin, Jonathan Braun, Weihua Cao, Liming Li, Dermot P.B. McGovern

## Abstract

**Introduction:** Coronavirus Disease 2019 (COVID-19) infection has been characterized by rapid spread and unusually large case clusters. It is important to have an estimate of the current state of COVID-19 epidemic in the U.S. to help develop informed public health strategies.

**Methods:** We estimated the potential scale of the COVID-19 epidemic (as of 03/01/2020) in the U.S. from cases ‘imported’ directly from Wuhan area. We used simulations based on transmission dynamics parameters estimated from previous studies and air traffic data from Wuhan to the U.S and deliberately built our model based on conservative assumptions. Detection and quarantine of individual COVID-19 cases in the U.S before 03/01/2020 were also taken into account. A SEIR model was used to simulate the growth of the number of infected individuals in Wuhan area and in the U.S.

**Results:** With the most likely model, we estimated that there would be 9,484 infected cases (90%CI 2,054-24,241) as of 03/01/2020 if no successful intervention procedure had been taken to reduce the transmissibility in unidentified cases. Assuming current preventive procedures have reduced 25% of the transmissibility in unidentified cases, the number of infected cases would be 1,043 (90%CI 107-2,474).

**Conclusion:** Our research indicates that, as of 03/01/2020., it is likely that there are already thousands of individuals in the US infected with SARS-CoV-2. Our model is dynamic and is available to the research community to further evaluate as the situation becomes clearer.

## Introduction

The Coronavirus Disease 2019 (COVID-19), an infectious disease caused by the Severe Acute Respiratory Syndrome Coronavirus 2(SARS-CoV-2)(1), originated in the central Chinese city of Wuhan(2). Following the onset of first reported COVID-19 patient on Dec 1^st^ 2019(3, 4), this disease has spread to over 60 countries in three months and has affected more than 90,000 people claiming more than 3,000 lives worldwide(5). As of the drafting of this manuscript (02/29/2020), COVID-19 outbreak had been reported in countries with large volumes of air traffic with China such as South Korea and Japan with thousands of people infected (6).

While the U.S. is one of the countries with the most air traffic to and from China, only a limited number of domestic COVID-19 cases (20 cases as of 02/29/2020) have been identified in the U.S. (7). Preemptively, the U.S. government took a number of steps to contain COVID-19, including travel alerts and travel bans, communication and advice to the general public and healthcare professionals, screening of incoming passengers, as well as case detection and contact tracing(8, 9). Despite these efforts, multiple COVID-19 cases with no relevant travel history or exposure to confirmed cases have recently been reported in the U.S., indicating possible community transmission of COVID-19(10, 11).

With the characteristics of COVID-19 including the potential of rapid spreading (12, 13) and unusually large case clusters(12, 14, 15), it would be crucial to have an estimate of the current state of the COVID-19 epidemic in the U.S. in order to develop informed public health strategies. Understanding the current state of COVID-19 can also be important for the general public to take informed personal and social preventive actions. Furthermore, developing and disseminating models to help understand the extent of COVID-19 infection will allow the research community and public health officials in other countries to further evaluate situations as the epidemic evolves worldwide.

In the current study, we estimated the scale of the COVID-19 epidemic in the U.S. based on transmission dynamics parameters estimated from previous studies and air traffic data between Wuhan and U.S., with the identification and quarantine of individual domestic COVID-19 case in the U.S. taken into account.

## Methods

### Assumptions

We have, by necessity, made a number of assumptions in modeling the transmission of COVID- 19. In most cases we believe that we have erred on the conservative side with our assumptions and have explicitly stated our assumptions along with our rationale so readers can infer their own opinion.

To simplify the calculation and consistent with our conservative approach, we have only estimated spreading of COVID-19 in the U.S. due to cases imported directly from the Wuhan area before the city lockdown by the Chinese government was implemented (01/23/2020). We have not included additional potential cases ‘imported’ through other parts of China or from other heavily affected countries such as South Korea, Italy or Iran(6). We have also assumed that with the lockdown of the Wuhan area, outbound traffic was completely shut down and so there would be no additional imported cases from the Wuhan area after 01/23/2020.

Similarly, we assumed that the imported cases were no longer spreading infection after they were diagnosed in the U.S., and that public health procedures in the Wuhan area did not change the parameters of the transmission model before 01/23/2020.

Several studies have estimated parameters for the transmission model of COVID-19, including basic reproduction number (*R*_*0*_), incubation period and serial interval (13, 16-26). These have largely been based on data from China or cases exported from China. Few data on transmission dynamics of COVID-19 in non-Asian populations are available. In the current investigation, we assumed that without intervention, the transmission parameters estimated from China would be the same in the U.S. population.

### Parameters and data source

Information on the confirmed domestic COVID-19 cases in the U.S. by 02/29/2020 (Table1) was obtained from CDC press releases or media briefings (11, 16, 27-33). Of the 20 cases, 8 (case #1- 5, #8-10) visited Wuhan before disease onset. Although travel dates were provided for only 2 of the 8 cases (case #1, returned from Wuhan on 01/15/2020 and case #2, returned from Wuhan on 01/13/2020), based on the assumption that there was a complete shutdown of the traffic in the Wuhan area, we assume that all 8 cases left Wuhan area before 01/23/2020.

Of the remaining 12 patients, two cases (case #7 and #11) were person-to-person transmissions. There was also 1 case with travel history to Beijing, 4 cases with unstated travel history and 5 cases with neither travel history to China nor previously known contact with confirmed cases. Taken together, this information suggested that there were at least 8 infected individuals imported from the Wuhan area before the lockdown.

Estimates of *R*_*0*_, which is the expected number of cases directly generated by one case in a population where all individuals are susceptible to infection(34), ranges from 2.1 to 3.5 for SARS-CoV-2 in previous studies (13, 16, 17, 19, 20, 22, 24, 25). In the current report we have only shown results for scenarios with *R*_*0*_ ranging from 2.1 to 2.5.

The incubation period of COVID-19 was set at 6 days(13, 21, 23) and the serial interval was set at 7.5 days(13, 26) based on previous publications. There have been reports on the possibility that COVID-19 cases can be infectious before disease onset(35, 36) and indication of asymptomatic individuals are able to transmit the virus to others(37, 38). However, there is no report on the latent period of COVID-19 which is defined as the interval between exposure and the onset of the period of communicability. We assumed a latent period of 2 days as there are reports that disease onset in some patients can be as early as 2 days after infection(13, 21). Infectious period, defined as the time during which an infected individual is infectious, was therefore calculated as:

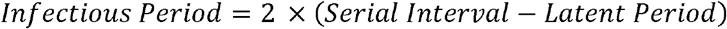

We adopted population parameters from the seminal work of Wu et al(19) assuming the following: a total population size in the Wuhan area of *N*_*w*_=19,000,000; a daily average number of international outbound air passengers *L*_*W,I*_=3,633; a daily average number of international inbound air passengers *L*_*I,W*_=3, 546; a daily number of domestic outbound traveler *L*_*W,C(t)*_=502,013; a daily number of average domestic inbound travelers *L*_*C,W(t)*_=487, 310 in the Wuhan area at time *t* before the Spring Festival travel season (01/10/2020); and an annual immigration after 01/10/2020 with daily numbers of domestic outbound and inbound travelers of *L*_*W,C(t)*_=717,226 and *L*_*C,W(t)*_=810,500 in the Wuhan area.

There were two air routes with direct flights from Wuhan Tianhe International Airport (WUH) to the U.S., one from WUH to San Francisco International Airport (SFO) and another new route (starting July 2019) from WUH to John F. Kennedy International Airport in New York (JFK). Air traffic data (Table 2, before September 2019) from WUH to the U.S. was obtained from the Bureau of Transportation Statistics (https://www.bts.gov/). The Air traffic from WUH to SFO in December 2019 and January 2020 was estimated using the traffic data of December 2018 and January 2019, respectively since volume of air traffic can be different in January with the Spring Festival travel season. Air traffic data from WUH to JFK was estimated using the traffic data of August 2019, as that is the only traffic data currently available for this new route.

**Table 1.** List of confirmed domestic COVID-19 cases in the U.S. as of 02/29/2020.

| State | Gender | Traveled history | Date_return | Date_diagnosis | Index |
| --- | --- | --- | --- | --- | --- |
| Washington | M | Wuhan | Jan 15. | Jan. 21 | 1 |
| Illinois | F | Wuhan | Jan.13 | Jan. 24 | 2 |
| California | Unknown | Wuhan | Unknown | Jan. 26 | 3 |
| California | Unknown | Wuhan | Unknown | Jan. 26 | 4 |
| Arizona | Unknown | Wuhan | Unknown | Jan.26 | 5 |
| Illinois | M | NO | - | Jan. 30 | 6 |
| California | M | Wuhan | Unknown | Jan. 31 | 7 |
| Massachusetts | Unknown | Wuhan | Unknown | Feb. 1 | 8 |
| California | F | Wuhan | Unknown | Feb. 2 | 9 |
| California | M | NO | - | Feb. 2 | 10 |
| California | F | Unknown | - | Feb. 2 | 11 |
| Wisconsin | F | Beijing | Unknown | Feb. 7 | 12 |
| California | F | Unknown | - | Feb. 21 | 13 |
| California | Unknown | Unknown | - | Feb. 21 | 14 |
| Washington | Unknown | Unknown | - | Feb. 26 | 15 |
| California | Unknown | NO | - | Feb. 28 | 16 |
| Oregon | Unknown | NO | - | Feb. 28 | 17 |
| Washington | Unknown | NO | - | Feb. 29 | 18 |
| Washington | Unknown | NO | - | Feb. 29 | 19 |
| Washington | Unknown | NO | - | Feb. 29 | 20 |

**Table 2.**
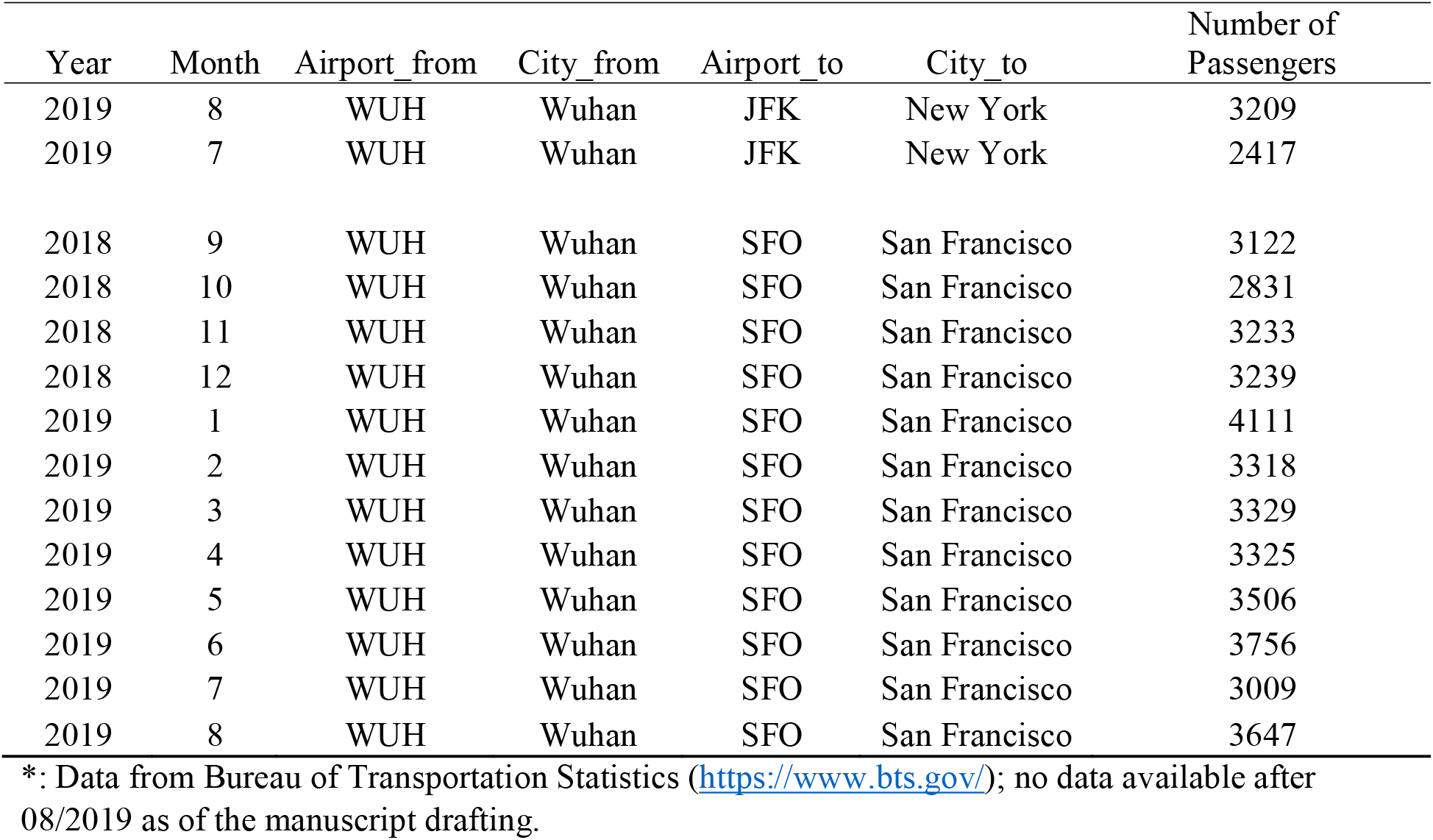
Air traffic statistics from Wuhan Tianhe Internation Airport (WUH) to United states*.

**Table 3.** Percentage of get 8 to 16 imported cases in simulations with different parameter settings.

| R0 | Z | Percentage of 8-16 imported cases |
| --- | --- | --- |
| 2.1 | 1.5 | 0.009 |
|  | 2 | 0.023 |
|  | 2.5 | 0.052 |
| 2.2 | 1.5 | 0.045 |
|  | 2 | 0.106 |
|  | 2.5 | 0.196 |
| 2.3 | 1.5 | 0.177 |
|  | 2 | 0.315 |
|  | 2.5 | 0.474 |
| 2.4 | 1.5 | 0.384 |
|  | 2 | 0.560 |
|  | 2.5 | 0.653 |
| 2.5 | 1.5 | 0.566 |
|  | 2 | 0.584 |
|  | 2.5 | 0.465 |

### Simulations

It was reported that the first COVID-19 case identified first exhibited symptoms on 12/01/2019(3, 4). We assume that the patient was infected on 11/25/2019, one incubation period earlier than the onset date, and 11/25/2019 was set as day 0 of the simulation.

Similar to Wu et al(19), We also simulated a constant zoonotic force to reflect the zoonotic effect (spreading of an infectious disease from non-human animals to humans) of the Huanan Seafood Market, which according to China CDC(3), was linked to 43 COVID-19 cases in Wuhan area and was shut down on 01/01/2020. We assumed the zoonotic force led to z=1.5/2/2.5 times of the observed infections linked to the market before 01/01/2020.

The spread of the disease in the Wuhan area was simulated using the SEIR model from Wu et al(19), with slight modification:

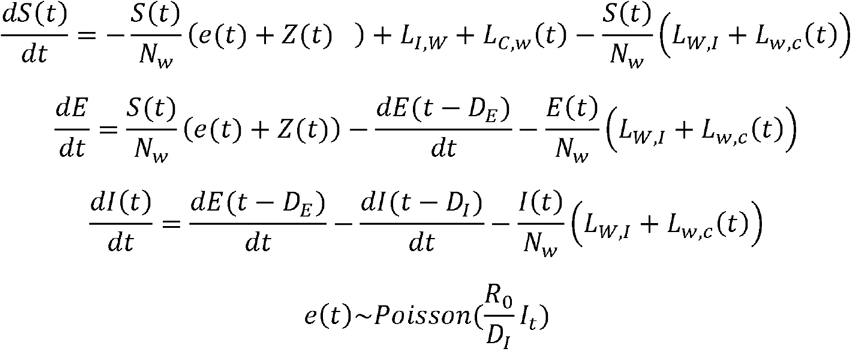

Where *S(t), E(t)* and *I(t)* represents the total number of susceptible, latent, and infectious individuals. *D*_*E*_ and *D*_*I*_ are length of latent and infectious period, respectively. For each day *t*, we randomly sampled from the simulated population to be the passengers traveling to the U.S. that day:

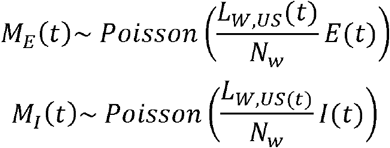

Where *M*_*E*_(t) and *M*_]_(t) are imported latent and infectious case via the air traffic from the Wuhan area to U.S., and L_W,US(t)_ represents the air traffic from WUH to U.S. at day *t*. Any infected individual in the sampled group, whether latent or infectious, were counted as imported cases and we assumed that the imported cases, when infectious, spread the infection until the date they were diagnosed in the U.S. based on a simplified SEIR model:

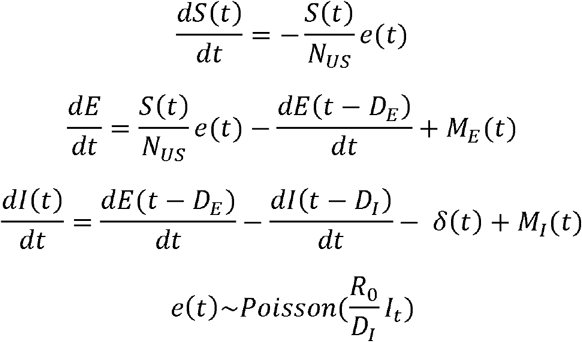

Here *N*_*US*_ was set to be 331 million assuming the whole population is susceptible to SARS-CoV-2 and o(t) represents real-life events (such as diagnosis and quarantine of an imported or 2^nd^ generation COVID-19 case) at time *t*.

In each simulation, disease spreading in the U.S. was simulated from the date the first imported case occurs until 03/01/2020, the target date of estimation. Considering the potential effect of the preventive procedures in the U.S., we simulated scenarios with different transmissibility of the un-identified cases in the U.S.

1000 simulations were performed for each different scenario. We monitored two key outcomes in each simulation: 1) the total number of imported cases from Wuhan area by 01/23/2020; 2) The total number of infected cases in the U.S. by 03/01/2020.

We assumed that U.S. CDC identified at least 50% of the imported cases, as recent investigation indicates that 46% infected cases of COVID-19 will not be detected by screening considering incubation period and proportion of asymptomatic cases(39). We thereby used the proportion of number of imported cases falling between 8-16 as a proxy to posterior likelihood of the model parameters. Conclusions were drawn based on the most plausible parameter settings of the simulations.

## Results

Figure 1 shows the distribution of imported COVID-19 cases from the Wuhan area to the U.S, with different R_0_ and zoonotic force z. A proxy of the posterior likelihood of different models, measured as proportion of imported cases falling between 8 and 16 cases by 01/23/2020 in 1000 simulations, are shown in Table3. Scenarios with R_0_ of 2.1-2.2 have a low percentage of observing 8-16 imported cases across different z value. The most plausible scenario is the setting with R_0_ of 2.4 and z of 2.5, with 8-16 imported cases observed in 65.3% simulations. Scenarios such as R_0_= 2.5 and z= 2 or 1.5 are slightly less plausible (56.6% and 58.4%, respectively).

**Figure 1:**
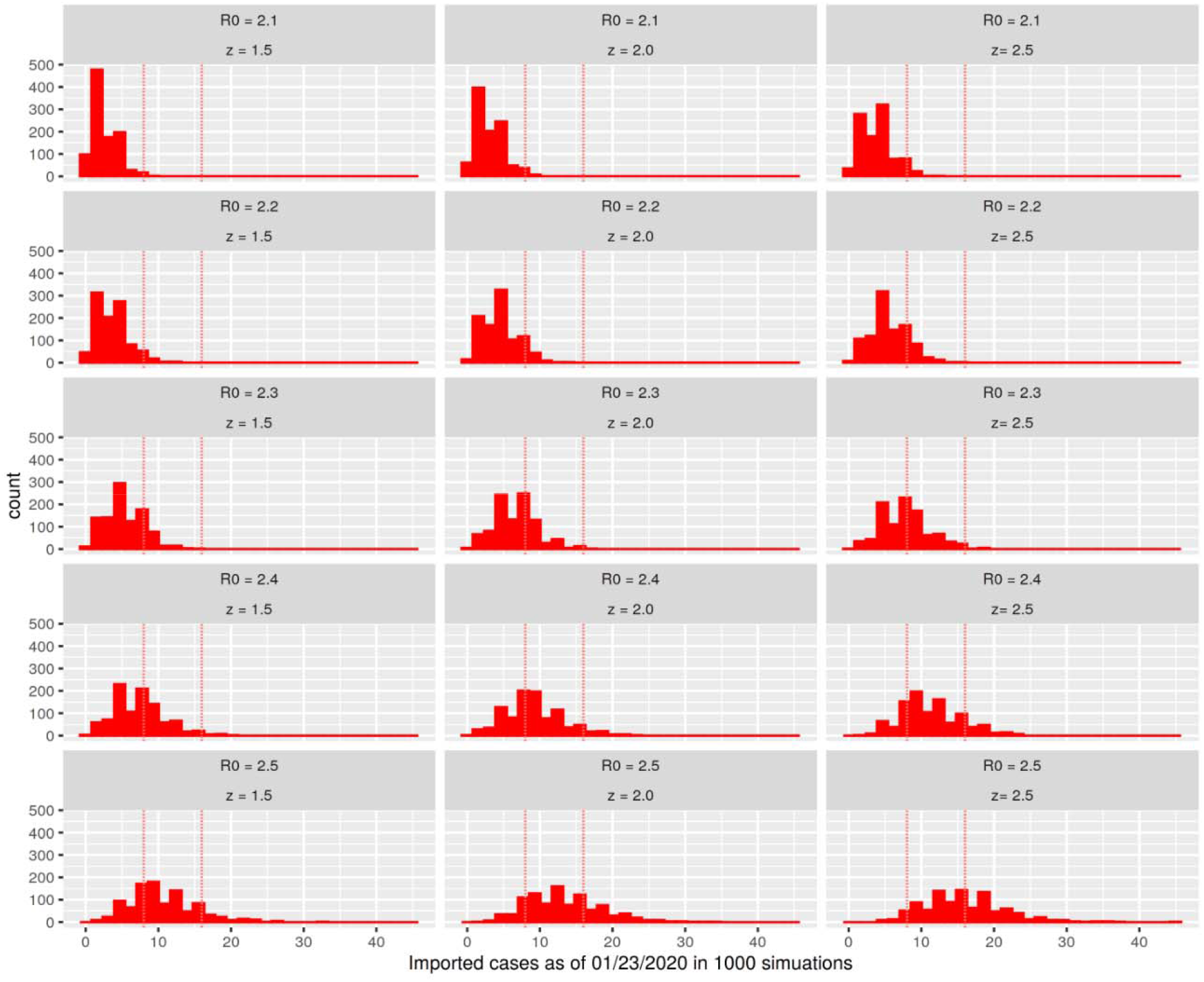
The distribution of imported cases from the Wuhan area to U.S. by 01/23/2020 in different simulated scenarios. Dotted lines represent the actual observed number of COVID-19 cases with Wuhan travel history (n=8) and an upper limit of 16 cases (assuming that U.S. CDC identified at least 50% of the imported cases) in the simulated scenario. *R*_*0*_: basic reproduction number; *z*: The zoonotic force that contributed to the spreading of disease in Wuhan area from 12/01/2020 to 01/01/2020, as a multiplier to the actual observed number (43) of COVID-19 cases linked to the Huanai Seafood Market.

Table 4 shows the estimated numbers of infected cases in the U.S. by 03/01/2020 across different scenarios if no intervention procedure had been successful in reducing the transmissibility in un-identified cases. In the most plausible scenario, with an R_0_ of 2.4 and z of 2.5, we estimate that there would be 9,484 infected cases (90%CI 2,054-24,241) in the U.S. by 03/01/2020. In other slightly less likely scenarios, e.g. with an R_0_ of 2.5 and z of 2, the number of infected cases would be 14,141 (90%CI:3,773-31,156).

**Table 4.** Estimated number of infected individuals in the U.S. by 03/01/2020, assuming no successful intervention procedure taken to reduce the transmissibility in un-identified cases.

| R0 | Z | Median | Quantile |  |
| --- | --- | --- | --- | --- |
|  |  |  | 5% | 95% |
| 2.3 | 1.5 | 4533 | 444 | 11316 |
|  | 2 | 5066 | 668 | 12294 |
|  | 2.5 | 5683 | 1198 | 13738 |
| 2.4 | 1.5 | 7975 | 1680 | 19370 |
|  | 2 | 9001 | 2299 | 20403 |
|  | 2.5 | 9484 | 2054 | 21241 |
| 2.5 | 1.5 | 13968 | 3971 | 30625 |
|  | 2 | 14141 | 3773 | 31156 |
|  | 2.5 | 16250 | 4187 | 33055 |

Table 5 shows the estimated number of infected cases if current preventive procedures reduced 25% of the transmissibility in un-identified cases. In the most plausible scenario (R_0_=2.4, z=2.5), there would be 1,013 infected cases (90%CI 107-2,474). In other slightly less likely scenarios (R_0_=2.5, z=1.5 or 2), the estimated numbers are 1,316 (90%CI 322-3,406) and 1,501(90%CI: 296-3,606), respectively.

**Table 5.** Estimated number of infected individuals in the U.S. by 03/01/2020, assuming successful intervention procedures reduced the transmissibility by 25% in un-identified cases.

| R0 | Z | Median | Quantile |  |
| --- | --- | --- | --- | --- |
|  |  |  | 5% | 95% |
| 2.3 | 1.5 | 531 | 13 | 1533 |
|  | 2 | 619 | 77 | 1628 |
|  | 2.5 | 587 | 21 | 1620 |
| 2.4 | 1.5 | 818 | 111 | 2043 |
|  | 2 | 914 | 101 | 2497 |
|  | 2.5 | 1013 | 107 | 2474 |
| 2.5 | 1.5 | 1316 | 322 | 3406 |
|  | 2 | 1501 | 296 | 3606 |
|  | 2.5 | 1697 | 432 | 3500 |

## Discussion

Here we estimated the potential scale of the COVID-19 epidemic in the U.S. exclusively from cases ‘imported’ directly from the Wuhan area using simulations across different possible scenarios. We identified the plausible scenarios by examining a proxy of the posterior probability of those models and estimated the number of infected cases in the U.S. by 03/01/2020 accordingly.

In previous studies, estimation of the epidemic of an infectious disease are often performed using mathematic inference with various transmission dynamic models(16, 19, 40, 41), usually in neat and elegant ways. Here we chose to use simulations based on the following considerations: 1) In many cases it would be difficult to consider irregular real-life events in mathematic inference, e.g. diagnosis and quarantine of an imported case at a given date. However, those events can be easily incorporated in simulations, as we have done here. 2) With the exponential spreading of the COVID-19, randomness in the early stages, e.g. whether one individual was infected or not on day 2, can have profound impact on the final estimates. This kind of randomness in theory can be calculated by summation over all possible scenarios but makes the inference cumbersome.

Simulation is a natural way to deal with this issue. 3) Comparing simulated results and real-life observation provides a way to identify optimal parameter settings. For example, in this study we have used the percentage of observing 8-16 imported cases as a proxy of the posterior probability of the simulated models. Although crude, this approach can give some indication on the most plausible models. We have made the code for the model freely available at github (link to be added) so that the research community and public officials in different countries can critically review and, we anticipate, refine the model based on both existing data and also as the epidemic resolves. This is the primary intention of our study.

In the simulations our approach has, deliberately, been very conservative. In our most plausible model, we used an R_0_ value (2.4) at the lower end of estimations from previous studies(13, 17, 20, 23). Our model has only studied the impact of travelers direct from the Wuhan area and not ‘indirect’ transfer from other parts of China (where tens of thousands of cases have been reported) or other countries. This makes our current estimation likely to be an underestimation of the true number of infected individuals in the U.S. This is clearly shown by the one confirmed imported COVID-19 case with travel history to Beijing, China (case #12).

Still, even with this overly conservative setting and an optimistic assumption that current procedures reduced the transmissibility of unidentified cases by 25%, the simulations estimate about one thousand infected cases as of 03/01 reside in the U.S. The real number of infected individuals is probably between one to ten thousand of infections based on the simulation across different scenarios. This suggests that the opportunity window to contain the epidemic of COVID-19 in its early stage is closing.

Our findings are consistent with recent research in which Bedford et al(42) demonstrated that based on the sequence similarity of different virus strains, the SARS-CoV-2 might have been spreading in community for about six weeks in the Seattle area. They also estimated that there were about 570 infected cases in Seattle area. Based on our simulations, there will probably be more case clusters similar to the situation in Seattle. Moreover, this echo of independent conclusions from molecular evidence and inference based on transmission dynamic models supports the validity of our research.

We cannot exclude the possibility that the simulation overestimates the number of U.S. infected individuals. There are almost no data on the transmission dynamic parameters of SARS-CoV-2 in non-Asian populations and our simulations assumed similar parameters as estimated based on the epidemic in China. It is possible that the transmission parameters are different in the U.S. as *R*_*0*_, the key parameter in the transmission dynamics model, can be viewed as a function of the likelihood of infection per contact between an infectious person and a suspectable person, and the contact rate(43). Factors that have the potential to influence the contact rate, like social organization and culture, might have an impact on *R*_*0*_(34). It is thus possible that differences in socio-economic, cultural and even environmental characteristics can lead to reduced transmissibility of the virus in the U. S. Moreover, the virus could have mutated to be less contagious and it is possible that current preventive procedures have reduced by more than the 25% transmissibility that we have assumed.

As reflected in our simulations, a key factor to mitigate the COVID-19 outbreak is to reduce the transmissibility of the unidentified cases. By reducing transmissibility by 25% in the early stage, the scope of an epidemic can be 10 times smaller. This will be a challenging task as a majority of COVID-19 cases are mild or even asymptomatic(15, 22, 23, 44, 45). Still this is possible either by aggressively mass screening for infected individuals, or by promoting social distancing, personal hygiene and restricting large-scale gatherings for occasions such as sporting events etc.

In conclusion, our model, based on a number of assumptions suggests that there are already thousands of individuals in the U.S. who have already been infected by SARS-CoV-2 as of 03/01/2020.

## Data Availability

All the data used in this manuscript are from public source.

